# Outbreak of P.3 (Theta) SARS-CoV-2 emerging variant of concern among service workers in Louisiana

**DOI:** 10.1101/2021.07.28.21259040

**Authors:** Rebecca Rose, David J. Nolan, Tessa M. LaFleur, Susanna L. Lamers

**Author notes:** Corresponding Author: Susanna L. Lamers, CEO, BioInfoExperts, LLC, 718 Bayou Ln, Thibodaux, LA, 70301.

## Abstract

In May, 2021, during routine oil and gas industrial quarantine/premobilization procedures, four individuals who recently arrived to Louisiana from the Philippines tested positive for SARS-CoV-2. Subsequent genomic analysis showed that all were infected with a Variant of Interest (P.3-Theta). This increases the number of known P.3 infections in the United States to eleven and highlights the importance of genomic surveillance within industries that are prone to rapidly spread the infection.

## MAIN TEXT

Many industries have implemented extensive safety measures to reduce workplace transmission risk during the COVID-19 pandemic. These policies have been imperative in the offshore oil and gas industry because rig workers travel from many states and internationally. The western and central Gulf of Mexico is one of the major offshore petroleum-producing areas, incorporating >1800 rigs and thousands of employees. To prevent COVID-19 outbreaks on offshore platforms, the industry has implemented, to varying degrees, strict pre-mobilization testing and quarantine policies.

Here, we report the detection of a rare Pango lineage [1], which infected four individuals who tested SARS-CoV-2 positive in May 2021 while quarantined in Louisiana, prior to deployment to an oil platform. These individuals were employed by a contractor to the oil and gas company, and all had recently traveled from the Philippines. Reconstruction of their timeline showed they tested negative in the Philippines, immediately prior to boarding a USA flight. Upon arrival in the USA, the workers went into a 14-day quarantine. They were retested after 9 days, at which point they were all qPCR+ and serology negative. None reported COVID-19 symptoms. The expected time interval between exposure and test positivity (3-4 days post-exposure for qPCR, >1 week post-exposure for serology), as well as the workers’ testing timeline (**Figure 1a**), suggest a recent exposure after their flight to the United States.

**Figure 1.**
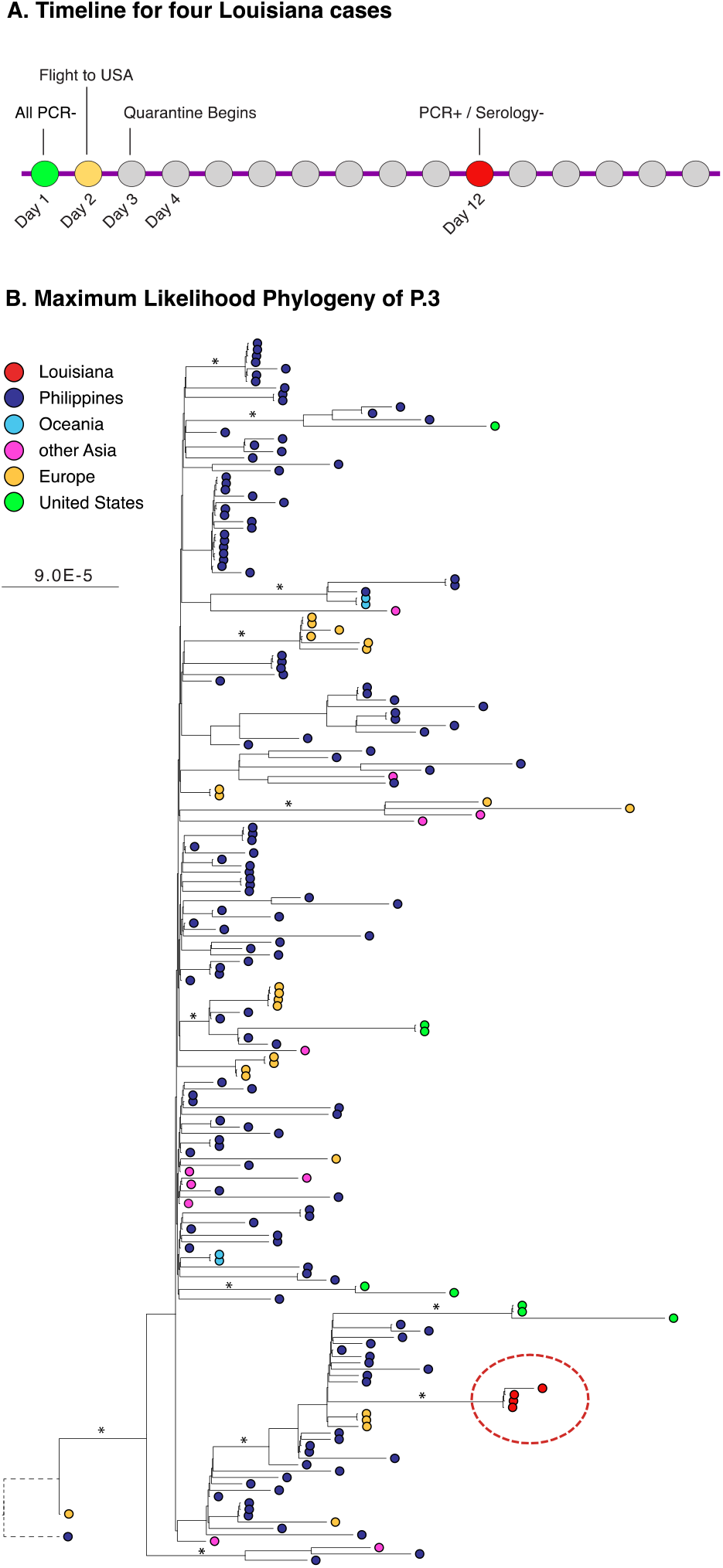
(A) Timeline for four Louisiana cases. Circles represent days. (B) Maximum likelihood phylogenetic tree of P.3. All sequences assigned to the P.3 lineage in GISAID as of 6/8/2021 (n=189) were included, as well as the four Louisiana sequences described. The tree is rooted using the Wuhan reference (EPI_ISL_402124). The circles at each tip represents a sequence, colored according to the region of sampling per the legend. Branches are scaled in substitutions per site.

To further understand the nature of the outbreak, the samples were sent to an independent laboratory (BioInfoExperts, LLC, Thibodaux, LA) for viral load and genomic evaluation (**Table 1**). Viral loads were quantified using ddPCR (BioRad). Sequencing was performed using the ARTIC protocol [2] on the Illumina MiSeq platform, followed by Pango lineage assignment [1] and mutation analysis using the FoxSeq software platform (https://bioinfox.com/foxseq/). All four samples were assigned to the Pango P.3. (Theta) lineage (GenBank Accession #: MZ368550 – MZ368553).

**Table 1.** Laboratory results for four Louisiana cases.

| Sample Name | rtPCR Cycles |  |  | Pre-mobilization |  | Viral Load |  |
| --- | --- | --- | --- | --- | --- | --- | --- |
|  | S | N | ORF1ab | PCR | Serology | N1 | N2 |
| BIE-120 | 16.44 | 16.685 | 16.286 | + | - | 20,000,000 | 20,000,000 |
| BIE-121 | 17.43 | 16.496 | 16.608 | + | - | 230,967 | 214,658 |
| BIE-122 | 11.469 | 12.289 | 10.811 | + | - | 55,109 | 54,942 |
| BIE-123 | 18.69 | 20.724 | 18.881 | + | - | 231,714 | 231,714 |

The P.3 lineage likely emerged from the Central Visayas region in the Philippines[3], the location of the earliest sampled sequence in this lineage in GISAID (January 2021). The majority of sequences assigned to this comparatively rare lineage are from the Philippines, although it has also been detected in the United States (n=8), Europe (n=24), and Oceania (n=4). The earliest identified P.3 infection in the United States was at the end of March in California (n=3), with additional cases detected in Washington (n=3), Virginia (n=1), and West Virginia (n=1).

To address a potential outbreak, we downloaded the masked alignment of all sequences submitted to GISAID (www.gisaid.org), extracted all sequences assigned to the P3 lineage (n=189), and inferred a maximum likelihood phylogenetic tree using IQTREE v2 [4] (**Figure 1b; Figure S1; Table S1**). The four Louisiana cases are clearly derived from a single origin and are separated from the most related P.3 sequences by a long branch, which likely resulted from the substantial time separating the Louisiana samples from the rest (2+ months). The sequences are part of a well-supported clade whose basal members are all from the Philippines. Elsewhere on the tree, well-supported clades of P.3 sequences from Europe, USA, Oceania, and/or Asia indicate multiple introductions to those locations. Interestingly, the most basal sequence of the entire P.3 clade (closest to the root of the tree) is from Europe, although it is separated from the rest of the P.3 tree by a long branch.

P.3 is a considered a Variant Under Investigation (VUI) by Public Health England (PHE) [5] and was formerly considered a Variant of Interest by the World Health Organization, as all P.3 viruses contain multiple concerning mutations shared with other Variants of Concern as denoted by PHE (VOCs; B.1.1.7, P.1, and B.1.351) in the spike protein, including E484K, N501Y, and P681H [6], which have been linked to increased transmissibility and immune escape [7, 8]. In addition, >95% of P.3. sequences exhibit spike protein mutations at D614G, H1101Y, E1092K, and V1176F. Two synonymous spike protein mutations also occur at positions 593G and 875S. The four new Louisiana sequences have a unique spike mutation at Q1180H, which was not observed in any of the other P.3 sequences. A three amino acid deletion (LGV141_143del) in the spike protein reported as common in this lineage [6] was not found in the four Louisiana P.3 sequences.

The identification of a novel Louisiana P.3 cluster is of particular concern given the suite of spike mutations, the relatively low vaccination rate in Louisiana (<40%; [9]), and the close working conditions on the oil rigs. Our results show that industrial monitoring for COVID-19 remains crucial. Fortunately, due the testing policies implemented, this particular variant was identified prior to deployment so that the workers continued quarantine until they were no longer considered a risk. Other employees have not contracted P.3 and are thought not to have been exposed. The study confirms that SARS-CoV-2 surveillance, especially in populations prone to viral spread, will remain important as the pandemic continues.

## Data Availability

Sequence data is available in GenBank, Accession #'s MZ368550 to MZ368553

## ACKNOWLEDGEMENTS

R.R., D.J.N., and S.L.L. are funded by National Science Foundation Small Business Innovation grants #1830867 and #2027424. The authors also thank Cheryl Metzler, CIH, and Valerie Murray, CIH.

## CONFLICT OF INTEREST

R.R., D.J.N., T.M.L, and S.L.L. are employed by BioInfoExperts LLC.

## ETHICS STATEMENT

A third party review from the WIRB-Copernicus Group (WCG IRB) concluded that this research was exempt from IRB review because it does not meet the definition of human subjects as defined in 45 CFR 46.102

## Notes

### Competing Interest Statement

All authors are employees by BioInfoExperts LLC

### Clinical Trial

Not a clinical trial

### Funding Statement

The project was supported in part by National Science Foundation Small Business Innovation grants # 1830867 and # 2027424

## References

1. Rambaut, A., et al., A dynamic nomenclature proposal for SARS-CoV-2 lineages to assist genomic epidemiology. Nat Microbiol, 2020. 5(11): p. 1403–1407.

2. Quick, J., et al., Multiplex PCR method for MinION and Illumina sequencing of Zika and other virus genomes directly from clinical samples. Nat Protoc, 2017. 12(6): p. 1261–1276.

3. Staff, C.P. DOH confirms new COVID-19 mutations in Central Visayas. 2021; Available from: https://www.cnnphilippines.com/news/2021/2/18/two-covid-mutations-of-concern-cebu-doh-central-visayas.html.

4. Minh, B.Q., et al., IQ-TREE 2: New Models and Efficient Methods for Phylogenetic Inference in the Genomic Era. Mol Biol Evol, 2020. 37(5): p. 1530–1534.

5. England, P.H. Variants of concern or under investigation: data up to 2 June 2021. 2021 June 9 2021]; Available from: https://www.gov.uk/government/publications/covid-19-variants-genomically-confirmed-case-numbers/variants-distribution-of-case-data-3-june-2021.

6. Tablizo, F.A., et al., Genome sequencing and analysis of an emergent SARS-CoV-2 variant characterized by multiple spike protein mutations detected from the Central Visayas Region of the Philippines. 2021, medRxiv.

7. Liu, Z., et al., Landscape analysis of escape variants identifies SARS-CoV-2 spike mutations that attenuate monoclonal and serum antibody neutralization. bioRxiv, 2020.

8. Greaney, A.J., et al., Comprehensive mapping of mutations in the SARS-CoV-2 receptor-binding domain that affect recognition by polyclonal human plasma antibodies. Cell Host Microbe, 2021. 29(3): p. 463-476.e6.

9. Clinic, M. Vaccine Tracker. 2021; Available from: https://www.mayoclinic.org/coronavirus-covid-19/vaccine-tracker.

