## Supplemental Table for "Outbreak of P.3 (Theta) SARS-CoV-2 emerging variant of concern among service workers in Louisiana"

Table S1. Metadata for GISAID sequences used in this study.

| gisaid_epi_isl | date | region | country | originating_lab | submitting_lab | authors |
| --- | --- | --- | --- | --- | --- | --- |
| EPI_ISL_1660475 | 2021-01-24 | Asia | Hong Kong | Department of Microbiology, The University of Hong Kong | The University of Hong Kong Department of Microbiology | Kelvin K.W. To et al |
| EPI_ISL_2296528 | 2021-03-25 | Asia | Hong Kong | Department of Health Technology and Informatics; The Hong Kong Polytechnic University | Department of Health Technology and Informatics; The Hong Kong Polytechnic University | Siu et al |
| EPI_ISL_2296527 | 2021-03-30 | Asia | Hong Kong | Department of Health Technology and Informatics; The Hong Kong Polytechnic University | Department of Health Technology and Informatics; The Hong Kong Polytechnic University | Siu et al |
| EPI_ISL_1198832 | 2021-02-25 | Asia | Japan | SARS-CoV-2 testing team; National Institute of Infectious Diseases | Pathogen Genomics Center; National Institute of Infectious Diseases | Tsuyoshi Sekizuka et al |
| EPI_ISL_1425715 | 2021-03-09 | Asia | Japan | SARS-CoV-2 testing team; National Institute of Infectious Diseases | Pathogen Genomics Center; National Institute of Infectious Diseases | Tsuyoshi Sekizuka et al |
| EPI_ISL_1425714 | 2021-03-11 | Asia | Japan | SARS-CoV-2 testing team; National Institute of Infectious Diseases | Pathogen Genomics Center; National Institute of Infectious Diseases | Tsuyoshi Sekizuka et al |
| EPI_ISL_1122458 | 2021-01-16 | Asia | Philippines | The Lord's Grace Medical and Industrial Clinic | Philippine Genome Center | Francis A. Tablizo et al |
| EPI_ISL_1213571 | 2021-01-17 | Asia | Philippines | Negros Oriental Provincial Hospital | Philippine Genome Center | Francis A. Tablizo et al |
| EPI_ISL_2156599 | 2021-01-20 | Asia | Philippines | Baguio General Hospital Medical Center (BGHMC) | Philippine Genome Center | Francis A. Tablizo et al |
| EPI_ISL_1122455 | 2021-01-23 | Asia | Philippines | Southern Philippines Medical Center | Philippine Genome Center | Francis A. Tablizo et al |
| EPI_ISL_1213568 | 2021-01-26 | Asia | Philippines | Lung Center of the Philippines (LCP) | Philippine Genome Center | Francis A. Tablizo et al |
| EPI_ISL_1122456 | 2021-01-28 | Asia | Philippines | Manila Doctors Hospital | Philippine Genome Center | Francis A. Tablizo et al |
| EPI_ISL_1122436 | 2021-01-31 | Asia | Philippines | Cebu TB Reference Laboratory | Philippine Genome Center | Francis A. Tablizo et al |
| EPI_ISL_1122451 | 2021-01-31 | Asia | Philippines | Cebu TB Reference Laboratory | Philippine Genome Center | Francis A. Tablizo et al |
| EPI_ISL_1122457 | 2021-02-01 | Asia | Philippines | Hi-Precision Diagnostics | Philippine Genome Center | Francis A. Tablizo et al |
| EPI_ISL_1122429 | 2021-02-01 | Asia | Philippines | Cebu TB Reference Laboratory | Philippine Genome Center | Francis A. Tablizo et al |
| EPI_ISL_1122430 | 2021-02-01 | Asia | Philippines | Cebu TB Reference Laboratory | Philippine Genome Center | Francis A. Tablizo et al |
| EPI_ISL_1122431 | 2021-02-01 | Asia | Philippines | Cebu TB Reference Laboratory | Philippine Genome Center | Francis A. Tablizo et al |
| EPI_ISL_1122432 | 2021-02-01 | Asia | Philippines | Cebu TB Reference Laboratory | Philippine Genome Center | Francis A. Tablizo et al |
| EPI_ISL_1122437 | 2021-02-01 | Asia | Philippines | Cebu TB Reference Laboratory | Philippine Genome Center | Francis A. Tablizo et al |
| EPI_ISL_1122438 | 2021-02-01 | Asia | Philippines | Cebu TB Reference Laboratory | Philippine Genome Center | Francis A. Tablizo et al |
| EPI_ISL_1122439 | 2021-02-01 | Asia | Philippines | Cebu TB Reference Laboratory | Philippine Genome Center | Francis A. Tablizo et al |
| EPI_ISL_1122440 | 2021-02-01 | Asia | Philippines | Cebu TB Reference Laboratory | Philippine Genome Center | Francis A. Tablizo et al |
| EPI_ISL_1122441 | 2021-02-01 | Asia | Philippines | Cebu TB Reference Laboratory | Philippine Genome Center | Francis A. Tablizo et al |
| EPI_ISL_1122442 | 2021-02-01 | Asia | Philippines | Cebu TB Reference Laboratory | Philippine Genome Center | Francis A. Tablizo et al |
| EPI_ISL_1122443 | 2021-02-01 | Asia | Philippines | Cebu TB Reference Laboratory | Philippine Genome Center | Francis A. Tablizo et al |
| EPI_ISL_1122444 | 2021-02-01 | Asia | Philippines | Cebu TB Reference Laboratory | Philippine Genome Center | Francis A. Tablizo et al |
| EPI_ISL_1122445 | 2021-02-01 | Asia | Philippines | Cebu TB Reference Laboratory | Philippine Genome Center | Francis A. Tablizo et al |
| EPI_ISL_1122446 | 2021-02-01 | Asia | Philippines | Cebu TB Reference Laboratory | Philippine Genome Center | Francis A. Tablizo et al |
| EPI_ISL_1122452 | 2021-02-01 | Asia | Philippines | Cebu TB Reference Laboratory | Philippine Genome Center | Francis A. Tablizo et al |
| EPI_ISL_1122453 | 2021-02-01 | Asia | Philippines | Cebu TB Reference Laboratory | Philippine Genome Center | Francis A. Tablizo et al |
| EPI_ISL_1122427 | 2021-02-01 | Asia | Philippines | Cebu TB Reference Laboratory | Philippine Genome Center | Francis A. Tablizo et al |
| EPI_ISL_1122454 | 2021-02-01 | Asia | Philippines | Cebu TB Reference Laboratory | Philippine Genome Center | Francis A. Tablizo et al |
| EPI_ISL_1213533 | 2021-02-01 | Asia | Philippines | Cebu TB Reference Laboratory | Philippine Genome Center | Francis A. Tablizo et al |
| EPI_ISL_1122433 | 2021-02-02 | Asia | Philippines | Cebu TB Reference Laboratory | Philippine Genome Center | Francis A. Tablizo et al |
| EPI_ISL_1122434 | 2021-02-02 | Asia | Philippines | Cebu TB Reference Laboratory | Philippine Genome Center | Francis A. Tablizo et al |
| EPI_ISL_1122435 | 2021-02-02 | Asia | Philippines | Cebu TB Reference Laboratory | Philippine Genome Center | Francis A. Tablizo et al |
| EPI_ISL_1122447 | 2021-02-02 | Asia | Philippines | Cebu TB Reference Laboratory | Philippine Genome Center | Francis A. Tablizo et al |
| EPI_ISL_1122448 | 2021-02-02 | Asia | Philippines | Cebu TB Reference Laboratory | Philippine Genome Center | Francis A. Tablizo et al |
| EPI_ISL_1122426 | 2021-02-02 | Asia | Philippines | Cebu TB Reference Laboratory | Philippine Genome Center | Francis A. Tablizo et al |
| EPI_ISL_1122449 | 2021-02-02 | Asia | Philippines | Cebu TB Reference Laboratory | Philippine Genome Center | Francis A. Tablizo et al |
| EPI_ISL_1122450 | 2021-02-02 | Asia | Philippines | Cebu TB Reference Laboratory | Philippine Genome Center | Francis A. Tablizo et al |
| EPI_ISL_1122428 | 2021-02-02 | Asia | Philippines | Cebu TB Reference Laboratory | Philippine Genome Center | Francis A. Tablizo et al |
| EPI_ISL_1219701 | 2021-02-03 | Asia | Philippines | The Lord's Grace Medical and Industrial Clinic | Philippine Genome Center | Francis A. Tablizo et al |
| EPI_ISL_1213583 | 2021-02-03 | Asia | Philippines | Teresita Jalandoni Provincial Hospital | Philippine Genome Center | Francis A. Tablizo et al |
| EPI_ISL_1213518 | 2021-02-03 | Asia | Philippines | Bohol Containerized PCR Laboratory | Philippine Genome Center | Francis A. Tablizo et al |
| EPI_ISL_1213509 | 2021-02-03 | Asia | Philippines | Bohol Containerized PCR Laboratory | Philippine Genome Center | Francis A. Tablizo et al |
| EPI_ISL_1213531 | 2021-02-03 | Asia | Philippines | Bohol Containerized PCR Laboratory | Philippine Genome Center | Francis A. Tablizo et al |
| EPI_ISL_1213521 | 2021-02-03 | Asia | Philippines | Bohol Containerized PCR Laboratory | Philippine Genome Center | Francis A. Tablizo et al |
| EPI_ISL_1213504 | 2021-02-03 | Asia | Philippines | Bohol Containerized PCR Laboratory | Philippine Genome Center | Francis A. Tablizo et al |
| EPI_ISL_1213524 | 2021-02-04 | Asia | Philippines | Bohol Containerized PCR Laboratory | Philippine Genome Center | Francis A. Tablizo et al |
| EPI_ISL_1213523 | 2021-02-04 | Asia | Philippines | Bohol Containerized PCR Laboratory | Philippine Genome Center | Francis A. Tablizo et al |
| EPI_ISL_1213561 | 2021-02-05 | Asia | Philippines | Cebu TB Reference Laboratory | Philippine Genome Center | Francis A. Tablizo et al |
| EPI_ISL_1213526 | 2021-02-05 | Asia | Philippines | Bohol Containerized PCR Laboratory | Philippine Genome Center | Francis A. Tablizo et al |
| EPI_ISL_1213516 | 2021-02-05 | Asia | Philippines | Bohol Containerized PCR Laboratory | Philippine Genome Center | Francis A. Tablizo et al |
| EPI_ISL_1213519 | 2021-02-05 | Asia | Philippines | Bohol Containerized PCR Laboratory | Philippine Genome Center | Francis A. Tablizo et al |
| EPI_ISL_1213530 | 2021-02-05 | Asia | Philippines | Bohol Containerized PCR Laboratory | Philippine Genome Center | Francis A. Tablizo et al |
| EPI_ISL_1213514 | 2021-02-05 | Asia | Philippines | Bohol Containerized PCR Laboratory | Philippine Genome Center | Francis A. Tablizo et al |
| EPI_ISL_1213569 | 2021-02-06 | Asia | Philippines | Manila Doctors Hospital | Philippine Genome Center | Francis A. Tablizo et al |
| EPI_ISL_1213535 | 2021-02-06 | Asia | Philippines | Cebu TB Reference Laboratory | Philippine Genome Center | Francis A. Tablizo et al |
| EPI_ISL_2091461 | 2021-02-06 | Asia | Philippines | Cebu TB Reference Laboratory | Philippine Genome Center | Francis A. Tablizo et al |
| EPI_ISL_1213556 | 2021-02-08 | Asia | Philippines | Cebu TB Reference Laboratory | Philippine Genome Center | Francis A. Tablizo et al |
| EPI_ISL_1213513 | 2021-02-08 | Asia | Philippines | Bohol Containerized PCR Laboratory | Philippine Genome Center | Francis A. Tablizo et al |
| EPI_ISL_1213511 | 2021-02-08 | Asia | Philippines | Bohol Containerized PCR Laboratory | Philippine Genome Center | Francis A. Tablizo et al |
| EPI_ISL_1213506 | 2021-02-08 | Asia | Philippines | Bohol Containerized PCR Laboratory | Philippine Genome Center | Francis A. Tablizo et al |
| EPI_ISL_1213528 | 2021-02-08 | Asia | Philippines | Bohol Containerized PCR Laboratory | Philippine Genome Center | Francis A. Tablizo et al |
| EPI_ISL_2091459 | 2021-02-08 | Asia | Philippines | Hi-Precision Diagnostic Center (QC) | Philippine Genome Center | Francis A. Tablizo et al |
| EPI_ISL_2105547 | 2021-02-08 | Asia | Philippines | Chinese General Hospital | Philippine Genome Center | Francis A. Tablizo et al |
| EPI_ISL_2156481 | 2021-02-08 | Asia | Philippines | Bohol Containerized PCR Laboratory | Philippine Genome Center | Francis A. Tablizo et al |
| EPI_ISL_1213539 | 2021-02-09 | Asia | Philippines | Cebu TB Reference Laboratory | Philippine Genome Center | Francis A. Tablizo et al |
| EPI_ISL_1213540 | 2021-02-09 | Asia | Philippines | Cebu TB Reference Laboratory | Philippine Genome Center | Francis A. Tablizo et al |
| EPI_ISL_1213546 | 2021-02-09 | Asia | Philippines | Cebu TB Reference Laboratory | Philippine Genome Center | Francis A. Tablizo et al |
| EPI_ISL_2156475 | 2021-02-09 | Asia | Philippines | Bohol Containerized PCR Laboratory | Philippine Genome Center | Francis A. Tablizo et al |
| EPI_ISL_1213551 | 2021-02-10 | Asia | Philippines | Cebu TB Reference Laboratory | Philippine Genome Center | Francis A. Tablizo et al |
| EPI_ISL_2156467 | 2021-02-10 | Asia | Philippines | Butuan Medical Center | Philippine Genome Center | Francis A. Tablizo et al |
| EPI_ISL_2156468 | 2021-02-10 | Asia | Philippines | Butuan Medical Center | Philippine Genome Center | Francis A. Tablizo et al |
| EPI_ISL_1213544 | 2021-02-11 | Asia | Philippines | Cebu TB Reference Laboratory | Philippine Genome Center | Francis A. Tablizo et al |
| EPI_ISL_1213508 | 2021-02-11 | Asia | Philippines | Bohol Containerized PCR Laboratory | Philippine Genome Center | Francis A. Tablizo et al |
| EPI_ISL_1213565 | 2021-02-13 | Asia | Philippines | Hi-Precision Diagnostic Center (QC) | Philippine Genome Center | Francis A. Tablizo et al |
| EPI_ISL_1213566 | 2021-02-13 | Asia | Philippines | Hi-Precision Diagnostic Center (QC) | Philippine Genome Center | Francis A. Tablizo et al |
| EPI_ISL_1213563 | 2021-02-13 | Asia | Philippines | Cotabato Regional and Medical Center | Philippine Genome Center | Francis A. Tablizo et al |
| EPI_ISL_1213581 | 2021-02-14 | Asia | Philippines | Prime Care Alpha Covid-19 Testing Laboratory | Philippine Genome Center | Francis A. Tablizo et al |
| EPI_ISL_1213549 | 2021-02-14 | Asia | Philippines | Cebu TB Reference Laboratory | Philippine Genome Center | Francis A. Tablizo et al |
| EPI_ISL_1213542 | 2021-02-14 | Asia | Philippines | Cebu TB Reference Laboratory | Philippine Genome Center | Francis A. Tablizo et al |
| EPI_ISL_2156391 | 2021-02-14 | Asia | Philippines | Philippine Red Cross - Port Area | Philippine Genome Center | Francis A. Tablizo et al |
| EPI_ISL_1213537 | 2021-02-15 | Asia | Philippines | Cebu TB Reference Laboratory | Philippine Genome Center | Francis A. Tablizo et al |
| EPI_ISL_1213558 | 2021-02-15 | Asia | Philippines | Cebu TB Reference Laboratory | Philippine Genome Center | Francis A. Tablizo et al |
| EPI_ISL_1213559 | 2021-02-15 | Asia | Philippines | Cebu TB Reference Laboratory | Philippine Genome Center | Francis A. Tablizo et al |
| EPI_ISL_1213554 | 2021-02-16 | Asia | Philippines | Cebu TB Reference Laboratory | Philippine Genome Center | Francis A. Tablizo et al |
| EPI_ISL_1213552 | 2021-02-16 | Asia | Philippines | Cebu TB Reference Laboratory | Philippine Genome Center | Francis A. Tablizo et al |
| EPI_ISL_1213547 | 2021-02-16 | Asia | Philippines | Cebu TB Reference Laboratory | Philippine Genome Center | Francis A. Tablizo et al |
| EPI_ISL_1219702 | 2021-02-16 | Asia | Philippines | Cebu TB Reference Laboratory | Philippine Genome Center | Francis A. Tablizo et al |
| EPI_ISL_2156359 | 2021-02-16 | Asia | Philippines | Research Institute for Tropical Medicine | Philippine Genome Center | Francis A. Tablizo et al |
| EPI_ISL_2156357 | 2021-02-16 | Asia | Philippines | Research Institute for Tropical Medicine | Philippine Genome Center | Francis A. Tablizo et al |
| EPI_ISL_2156354 | 2021-02-16 | Asia | Philippines | Research Institute for Tropical Medicine | Philippine Genome Center | Francis A. Tablizo et al |
| EPI_ISL_1213588 | 2021-02-18 | Asia | Philippines | The Medical City - Ortigas | Philippine Genome Center | Francis A. Tablizo et al |
| EPI_ISL_1213578 | 2021-02-18 | Asia | Philippines | Philippine Red Cross - Port Area | Philippine Genome Center | Francis A. Tablizo et al |
| EPI_ISL_1213576 | 2021-02-19 | Asia | Philippines | PHILIPPINE AIRPORT DIAGNOSTIC LABORATORY | Philippine Genome Center | Francis A. Tablizo et al |
| EPI_ISL_1213580 | 2021-02-20 | Asia | Philippines | Philippine Red Cross Logistics and Multipurpose Center | Philippine Genome Center | Francis A. Tablizo et al |
| EPI_ISL_2156294 | 2021-02-21 | Asia | Philippines | Philippine Red Cross Logistics and Multipurpose Center | Philippine Genome Center | Francis A. Tablizo et al |
| EPI_ISL_1213574 | 2021-02-22 | Asia | Philippines | PHILIPPINE AIRPORT DIAGNOSTIC LABORATORY | Philippine Genome Center | Francis A. Tablizo et al |
| EPI_ISL_2156284 | 2021-02-22 | Asia | Philippines | PHILIPPINE AIRPORT DIAGNOSTIC LABORATORY | Philippine Genome Center | Francis A. Tablizo et al |
| EPI_ISL_1213586 | 2021-02-23 | Asia | Philippines | The Lord's Grace Medical and Industrial Clinic | Philippine Genome Center | Francis A. Tablizo et al |
| EPI_ISL_2156264 | 2021-02-23 | Asia | Philippines | THE LORD'S GRACE MEDICAL AND INDUSTRIAL CLINIC | Philippine Genome Center | Francis A. Tablizo et al |
| EPI_ISL_2156263 | 2021-02-23 | Asia | Philippines | THE LORD'S GRACE MEDICAL AND INDUSTRIAL CLINIC | Philippine Genome Center | Francis A. Tablizo et al |
| EPI_ISL_1213585 | 2021-02-24 | Asia | Philippines | The Lord's Grace Medical and Industrial Clinic | Philippine Genome Center | Francis A. Tablizo et al |
| EPI_ISL_2188075 | 2021-03-01 | Asia | Philippines | Philippine Red Cross - Port Area | Philippine Genome Center | Francis A. Tablizo et al |
| EPI_ISL_2171237 | 2021-03-05 | Asia | Philippines | Philippine Red Cross - Port Area | Philippine Genome Center | Francis A. Tablizo et al |
| EPI_ISL_2171174 | 2021-03-05 | Asia | Philippines | Philippine Red Cross - Port Area | Philippine Genome Center | Francis A. Tablizo et al |
| EPI_ISL_2171239 | 2021-03-05 | Asia | Philippines | Philippine Red Cross - Port Area | Philippine Genome Center | Francis A. Tablizo et al |
| EPI_ISL_2188848 | 2021-03-05 | Asia | Philippines | Research Institute for Tropical Medicine | Philippine Genome Center | Francis A. Tablizo et al |
| EPI_ISL_2188756 | 2021-03-05 | Asia | Philippines | Research Institute for Tropical Medicine | Philippine Genome Center | Francis A. Tablizo et al |
| EPI_ISL_2188812 | 2021-03-06 | Asia | Philippines | Baguio General Hospital Medical Center (BGHMC) | Philippine Genome Center | Francis A. Tablizo et al |
| EPI_ISL_2171098 | 2021-03-07 | Asia | Philippines | National Kidney and Transplant Institute (NKTI) | Philippine Genome Center | Francis A. Tablizo et al |
| EPI_ISL_2171206 | 2021-03-08 | Asia | Philippines | South Super Highway Molecular Diagnostic Laboratory | Philippine Genome Center | Francis A. Tablizo et al |
| EPI_ISL_2188697 | 2021-03-08 | Asia | Philippines | Philippine Red Cross Logistics and Multipurpose Center | Philippine Genome Center | Francis A. Tablizo et al |
| EPI_ISL_2171148 | 2021-03-08 | Asia | Philippines | Philippine Red Cross - Port Area | Philippine Genome Center | Francis A. Tablizo et al |
| EPI_ISL_2188079 | 2021-03-08 | Asia | Philippines | Research Institute for Tropical Medicine | Philippine Genome Center | Francis A. Tablizo et al |
| EPI_ISL_2188203 | 2021-03-10 | Asia | Philippines | The Medical City -√Ç Ortigas | Philippine Genome Center | Francis A. Tablizo et al |
| EPI_ISL_2188206 | 2021-03-12 | Asia | Philippines | Laguna Provincial Hospital - San Pablo District Hospital | Philippine Genome Center | Francis A. Tablizo et al |
| EPI_ISL_2188699 | 2021-03-12 | Asia | Philippines | Philippine Red Cross - Port Area | Philippine Genome Center | Francis A. Tablizo et al |
| EPI_ISL_2188811 | 2021-03-12 | Asia | Philippines | Philippine Red Cross - Port Area | Philippine Genome Center | Francis A. Tablizo et al |
| EPI_ISL_2188167 | 2021-03-13 | Asia | Philippines | The Medical City -√Ç Ortigas | Philippine Genome Center | Francis A. Tablizo et al |
| EPI_ISL_2188826 | 2021-03-13 | Asia | Philippines | Philippine Red Cross - Port Area | Philippine Genome Center | Francis A. Tablizo et al |
| EPI_ISL_2188103 | 2021-03-14 | Asia | Philippines | University of the Philippines National Institutes of Health (UP NIH) | Philippine Genome Center | Francis A. Tablizo et al |
| EPI_ISL_2188853 | 2021-03-15 | Asia | Philippines | Philippine Red Cross - Port Area | Philippine Genome Center | Francis A. Tablizo et al |
| EPI_ISL_2188938 | 2021-03-18 | Asia | Philippines | Philippine Red Cross - Port Area | Philippine Genome Center | Francis A. Tablizo et al |
| EPI_ISL_2188892 | 2021-03-18 | Asia | Philippines | Philippine Red Cross - Port Area | Philippine Genome Center | Francis A. Tablizo et al |
| EPI_ISL_2189194 | 2021-03-18 | Asia | Philippines | Philippine Red Cross - Port Area | Philippine Genome Center | Francis A. Tablizo et al |
| EPI_ISL_2189380 | 2021-03-18 | Asia | Philippines | Butuan Medical Center | Philippine Genome Center | Francis A. Tablizo et al |
| EPI_ISL_2189095 | 2021-03-19 | Asia | Philippines | Philippine Red Cross - National Blood Center | Philippine Genome Center | Francis A. Tablizo et al |
| EPI_ISL_2189124 | 2021-03-19 | Asia | Philippines | Philippine Red Cross - Port Area | Philippine Genome Center | Francis A. Tablizo et al |
| EPI_ISL_2189046 | 2021-03-19 | Asia | Philippines | Butuan Medical Center | Philippine Genome Center | Francis A. Tablizo et al |
| EPI_ISL_2189296 | 2021-03-19 | Asia | Philippines | Butuan Medical Center | Philippine Genome Center | Francis A. Tablizo et al |
| EPI_ISL_2189233 | 2021-03-19 | Asia | Philippines | Butuan Medical Center | Philippine Genome Center | Francis A. Tablizo et al |
| EPI_ISL_2189273 | 2021-03-19 | Asia | Philippines | Butuan Medical Center | Philippine Genome Center | Francis A. Tablizo et al |
| EPI_ISL_2188870 | 2021-03-19 | Asia | Philippines | Butuan Medical Center | Philippine Genome Center | Francis A. Tablizo et al |
| EPI_ISL_2189035 | 2021-03-20 | Asia | Philippines | Philippine Red Cross - Port Area | Philippine Genome Center | Francis A. Tablizo et al |
| EPI_ISL_2188906 | 2021-03-21 | Asia | Philippines | Butuan Medical Center | Philippine Genome Center | Francis A. Tablizo et al |
| EPI_ISL_2189252 | 2021-03-21 | Asia | Philippines | Butuan Medical Center | Philippine Genome Center | Francis A. Tablizo et al |
| EPI_ISL_2189493 | 2021-03-21 | Asia | Philippines | Butuan Medical Center | Philippine Genome Center | Francis A. Tablizo et al |
| EPI_ISL_2189441 | 2021-03-22 | Asia | Philippines | Mary Mediatrix Medical Center | Philippine Genome Center | Francis A. Tablizo et al |
| EPI_ISL_2189408 | 2021-03-22 | Asia | Philippines | The Medical City | Philippine Genome Center | Francis A. Tablizo et al |
| EPI_ISL_2189350 | 2021-03-22 | Asia | Philippines | Northern Mindanao TB Regional Center | Philippine Genome Center | Francis A. Tablizo et al |
| EPI_ISL_2189187 | 2021-03-22 | Asia | Philippines | Northern Mindanao TB Regional Center | Philippine Genome Center | Francis A. Tablizo et al |
| EPI_ISL_2189152 | 2021-03-22 | Asia | Philippines | Northern Mindanao TB Regional Center | Philippine Genome Center | Francis A. Tablizo et al |
| EPI_ISL_2189463 | 2021-03-24 | Asia | Philippines | The Lord's Grace Medical and Industrial Clinic | Philippine Genome Center | Francis A. Tablizo et al |
| EPI_ISL_2189090 | 2021-03-25 | Asia | Philippines | The Lord's Grace Medical and Industrial Clinic | Philippine Genome Center | Francis A. Tablizo et al |
| EPI_ISL_2253110 | 2021-05-13 | Asia | Philippines | SARS-CoV-2 testing team; National Institute of Infectious Diseases | Pathogen Genomics Center; National Institute of Infectious Diseases | Tsuyoshi Sekizuka et al |
| EPI_ISL_1652091 | 2021-04-13 | Asia | Singapore | National Public Health Laboratory; National Centre for Infectious Diseases | National Public Health Laboratory; National Centre for Infectious Diseases | Tze Minn Mak et al |
| EPI_ISL_2349790 | 2021-05-03 | Asia | Singapore | National Public Health Laboratory; National Centre for Infectious Diseases | National Public Health Laboratory; National Centre for Infectious Diseases | Tze Minn Mak et al |
| EPI_ISL_1647347 | 2021-03-07 | Asia | South Korea | Division of Emerging Infectious Diseases; Bureau of Infectious Diseases Diagnosis Control; Korea Disease Control and Prevention Agency | Division of Emerging Infectious Diseases; Bureau of Infectious Diseases Diagnosis Control; Korea Disease Control and Prevention Agency | Ae Kyung Park et al |
| EPI_ISL_1153668 | 2021-02-11 | Europe | Germany | MVZ Labor Dr. Fenner und Kollegen (Standort Hamburg) | Robert Koch Institute | ? |
| EPI_ISL_1354960 | 2021-02-27 | Europe | Germany | Universit√§tsklinikum Heidelberg | Robert Koch Institute | ? |
| EPI_ISL_1354944 | 2021-03-01 | Europe | Germany | Universit√§tsklinikum Heidelberg | Robert Koch Institute | ? |
| EPI_ISL_1354990 | 2021-03-01 | Europe | Germany | Universit√§tsklinikum Heidelberg | Robert Koch Institute | ? |
| EPI_ISL_1570318 | 2021-03-19 | Europe | Germany | Universit√É¬§tsklinikum Heidelberg | Robert Koch Institute | ? |
| EPI_ISL_1570731 | 2021-03-23 | Europe | Germany | MVZ Labor Dr. Limbach & Kollegen GbR | Robert Koch Institute | ? |
| EPI_ISL_1571348 | 2021-03-28 | Europe | Germany | Eurofins LifeCodexx GmbH | Robert Koch Institute | ? |
| EPI_ISL_1643403 | 2021-03-29 | Europe | Germany | MVZ Labor Dr. Limbach & Kollegen GbR | Robert Koch Institute | ? |
| EPI_ISL_1722848 | 2021-04-04 | Europe | Germany | Universit√É¬§tsklinikum Heidelberg | Robert Koch Institute | ? |
| EPI_ISL_1597203 | 2021-02-27 | Europe | Netherlands | Dutch COVID-19 response team | National Institute for Public Health and the Environment (RIVM) | Adam Meijer et al |
| EPI_ISL_1792918 | 2021-03-20 | Europe | Netherlands | Dutch COVID-19 response team | National Institute for Public Health and the Environment (RIVM) | Adam Meijer et al |
| EPI_ISL_1522133 | 2021-03-22 | Europe | Netherlands | Dutch COVID-19 response team | National Institute for Public Health and the Environment (RIVM) | Adam Meijer et al |
| EPI_ISL_1522134 | 2021-03-22 | Europe | Netherlands | Dutch COVID-19 response team | National Institute for Public Health and the Environment (RIVM) | Adam Meijer et al |
| EPI_ISL_1792919 | 2021-03-22 | Europe | Netherlands | Dutch COVID-19 response team | National Institute for Public Health and the Environment (RIVM) | Adam Meijer et al |
| EPI_ISL_1597109 | 2021-03-29 | Europe | Netherlands | Dutch COVID-19 response team | National Institute for Public Health and the Environment (RIVM) | Adam Meijer et al |
| EPI_ISL_1323166 | 2021-02-01 | Europe | Norway | Department of Medical Microbiology; Molde Hospital | Norwegian Institute of Public Health; Department of Virology | Kathrine Stene-Johansen et al |
| EPI_ISL_1073934 | 2021-02-05 | Europe | Norway | Department of Medical Microbiology; Molde Hospital | Norwegian Institute of Public Health; Department of Virology | Kathrine Stene-Johansen et al |
| EPI_ISL_1256377 | 2021-03-01 | Europe | United Kingdom | Lighthouse Lab in Cambridge | Wellcome Sanger Institute for the COVID-19 Genomics UK (COG-UK) Consortium | Rob Howes et al |
| EPI_ISL_1256430 | 2021-03-01 | Europe | United Kingdom | Lighthouse Lab in Cambridge | Wellcome Sanger Institute for the COVID-19 Genomics UK (COG-UK) Consortium | Rob Howes et al |
| EPI_ISL_1244897 | 2021-03-03 | Europe | United Kingdom | Lighthouse Lab in Cambridge | Wellcome Sanger Institute for the COVID-19 Genomics UK (COG-UK) Consortium | Rob Howes et al |
| EPI_ISL_1333537 | 2021-03-03 | Europe | United Kingdom | Lighthouse Lab in Cambridge | Wellcome Sanger Institute for the COVID-19 Genomics UK (COG-UK) Consortium | Rob Howes et al |
| EPI_ISL_1275649 | 2021-03-05 | Europe | United Kingdom | Lighthouse Lab in Cambridge | Wellcome Sanger Institute for the COVID-19 Genomics UK (COG-UK) Consortium | Rob Howes et al |
| EPI_ISL_1986837 | 2021-04-23 | Europe | United Kingdom | Lighthouse Lab in Milton Keynes | Wellcome Sanger Institute for the COVID-19 Genomics UK (COG-UK) Consortium | The Lighthouse Lab in Milton Keynes et al |
| EPI_ISL_2352773 | 2021-05-25 | Europe | United Kingdom | Lighthouse Lab in Milton Keynes | Wellcome Sanger Institute for the COVID-19 Genomics UK (COG-UK) Consortium | The Lighthouse Lab in Milton Keynes et al |
| EPI_ISL_1525595 | 2021-03-25 | North America | USA | Fulgent Genetics | Centers for Disease Control and Prevention Division of Viral Diseases | Dakota Howard et al |
| EPI_ISL_1614114 | 2021-04-01 | North America | USA | Fulgent Genetics | Centers for Disease Control and Prevention Division of Viral Diseases | Dakota Howard et al |
| EPI_ISL_2229455 | 2021-04-29 | North America | USA | Virginia Division of Consolidated Laboratory Services | Virginia Division of Consolidated Laboratory Services | Virginia DCLS et al |
| EPI_ISL_2104833 | 2021-05-03 | North America | USA | UW Virology Lab | UW Virology Lab | Pavitra Roychoudhury et al |
| EPI_ISL_2161918 | 2021-05-03 | North America | USA | UW Virology Lab | UW Virology Lab | Pavitra Roychoudhury et al |
| EPI_ISL_2181026 | 2021-05-10 | North America | USA | Aegis Sciences Corporation | Centers for Disease Control and Prevention Division of Viral Diseases | Dakota Howard et al |
| EPI_ISL_2333059 | 2021-05-12 | North America | USA | UW Virology Lab | UW Virology Lab | Pavitra Roychoudhury et al |
| EPI_ISL_2370650 | 2021-05-23 | North America | USA | Aegis Sciences Corporation | Centers for Disease Control and Prevention Division of Viral Diseases | Dakota Howard et al |
| EPI_ISL_1273082 | 2021-03-15 | Oceania | Australia | Public Health Virology-Forensic and Scientific Services (PHV-FSS) | Public Health Virology-Forensic and Scientific Services (PHV-FSS) | Son Nguyen et al |
| EPI_ISL_1306121 | 2021-03-16 | Oceania | Australia | Public Health Virology-Forensic and Scientific Services (PHV-FSS) | Public Health Virology-Forensic and Scientific Services (PHV-FSS) | Son Nguyen et al |
| EPI_ISL_1315326 | 2021-03-13 | Oceania | New Zealand | LabPLUS | Institute of Environmental Science and Research (ESR) | Rachel Boyle et al |
| EPI_ISL_1469105 | 2021-03-19 | Oceania | New Zealand | Middlemore Hospital | Institute of Environmental Science and Research (ESR) | Rachel Boyle et al |
