## Supplementary figures and images for "Outbreak of P.3 (Theta) SARS-CoV-2 emerging variant of concern among service workers in Louisiana"

### Supplemental Figure

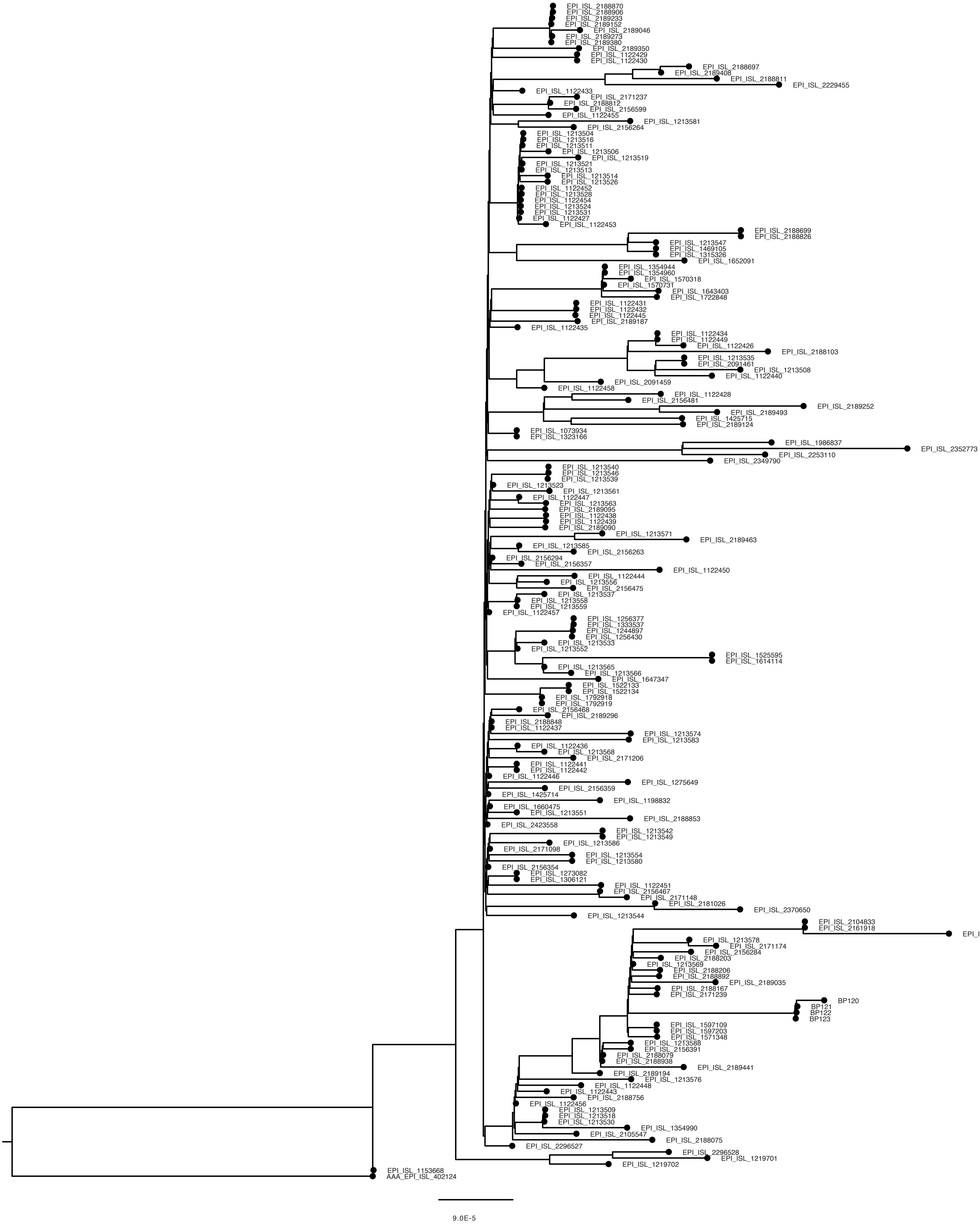
